# Lung cancer and Risk of Cardiovascular Mortality

**DOI:** 10.1101/2024.12.06.24318630

**Authors:** Chengshi Wang, Zhu Wang, Jing Yang, Songbo Zhang, Purong Zhang, Ye Yang

**Author notes:** These authors have contributed equally to this work (Chengshi Wang, Zhu Wang, Jing Yang). CORRESPONDENCE Purong Zhang Ye Yang.

## Abstract

**Purpose:** To investigate the cardiovascular mortality risk among lung cancer patients compared to the general population.

**Methods:** Using data from the National Cancer Institute’s Surveillance, Epidemiology, and End Results program, we conducted a population-based cohort study including 278,418 lung cancer patients over 30 years old between January 1, 1990, and December 31, 2020 as well as the general population. Poisson regression was employed to calculate incidence rate ratios (IRRs) for cardiovascular mortality.

**Results:** Patients exhibited a significantly higher incidence rate ratio (IRR) of cardiovascular mortality risk compared to the general population [IRR 1.74, 95% confidence interval (CI) 1.71-1.77]. The risk was most pronounced in patients aged 30-79 years (IRR 2.61, 95% CI 2.55-2.66), peaking at ages 30-34 (IRR 48.93, 95% CI 21.98-108.92). Elevated cardiovascular mortality risks were observed across all subgroups, including diseases of the heart (IRR 1.79, 95% CI 1.75-1.82), cerebrovascular diseases (IRR 1.52, 95% CI 1.45-1.59), and other cardiovascular diseases (IRR 1.78, 95% CI 1.67-1.90). The first month post-diagnosis presented the highest risk for ages 30-79 (IRR 12.08, 95% CI 11.49-12.70) and ≥80 years (IRR 4.03, 95% CI 3.70-4.39). Clinical characteristics significantly modified cardiovascular mortality.

**Conclusions:** Integrating cardiovascular disease monitoring and proactive management into lung cancer treatment protocols is essential to the improvement of overall survival and quality of life for lung cancer patients, particularly those who were young or with advanced tumor stage.

## Introduction

Lung cancer is a leading cause of cancer-related mortality worldwide, responsible for approximately 1.8 million deaths annually^1^. Despite advances in early detection, surgical techniques, and systemic therapies, the prognosis for lung cancer patients remains poor, with a five-year survival rate of approximately 20%. Lung cancer is often diagnosed at an advanced stage, complicating treatment and contributing to high mortality rates^2^.

The proportion of cardiovascular disease (CVD) deaths, leading cause of non-cancer death, is accounted for approximately 5%-10% of lung cancer patients^3–5^. Lung cancer and CVD share common risk factors such as smoking, age, obesity, hypertension, diabetes, sedentary behavior and also oxidative stress ^6–8^. These shared risk factors contribute to an increased baseline risk of cardiovascular events and cancer mortality in lung cancer patients^9^. Then, the treatments for lung cancer, including chemotherapy, radiotherapy, and targeted therapies, have cardiotoxic effects. Radiotherapy is associated with an increased risk of heart disease death in patients with lung cancer^10^. Platinum and immune checkpoint inhibitors, commonly used in lung cancer treatment, are well-known for their cardiotoxicity, leading to heart failure and other cardiovascular complications^11^. Newer targeted therapies, such as tyrosine kinase inhibitors, also pose risks for cardiovascular health^12^. Previous studies yielded the excess risk of CVD death increased in specific lung cancer patients (e.g., higher age, male gender, squamous cell carcinoma, left-side laterality)^3,9,13^. The cumulative burden of cancer treatment and the underlying cardiovascular risk factors necessitates a deeper understanding of cardiovascular mortality in this patient population. In addition, the inflammatory response induced by both cancer and its treatment plays a crucial role in CVD^14,15^. Moreover, the psychological stress associated with a lung cancer diagnosis can exacerbate risk of cardiovascular mortality^16^. Stress-related mechanisms, including increased sympathetic nervous system activity and elevated catecholamine levels, would precipitate cardiovascular events^17^. Lung cancer patients are subjected to a multifaceted absolute risk profile that significantly elevates the risk of cardiovascular mortality. However, there is limited comprehensive study quantifying relative risk of cardiovascular mortality in lung cancer patients compared with the general population.

The Surveillance, Epidemiology, and End Results (SEER) program provides a robust database that enables the examination of long-term trends and outcomes in individuals. By establishing a population-based cohort study including lung cancer patients and the general population, our study aims to identify age at follow-up, time since lung cancer diagnosis, demographic and clinical factors that influence the relative risk of CVD mortality when compared with general population.

## Material and methods

### Patients and Methods

We conducted a retrospective cohort study including patients over 30 years old who were diagnosed as first primary lung cancer during January 1, 1990 to December 31, 2020 in SEER database. We incorporated 53,324,676 per 100 person-years from the general population during 1990-2020 in the U.S. We also selected 448,794 patients who were diagnosed as first primary lung cancer by pathological confirmation from 1990 to 2020. We excluded patients who were diagnosed without pathological confirmation (N=48,598), whose first primary tumor was not malignancy (N=50), without county information. (N=119,217) or whose follow-up information was not calculated (N=827), without race report (N=486), with cancer in situ(N=1), whose malignancy was not carcinoma(N=747), younger than 30 years old(N=450). Finally, we included 278,418 lung cancer patients.

### Certification of CVD deaths and follow-up

CVD Death was defined as primary outcome. We used the International Classification of Diseases codes (Table S1) to confirm the death from CVD ^18^,which was furtherly stratified, as disease of the heart, cerebrovascular disease, or other CVD. Both patients and population follow-up were conducted regularly by differential hospital- and population-based registries. To guarantee the adequate follow-up visit, personal follow-up was also periodically visited for those who were considered lost to contact.

### Variables

Demographic Information on the patients and general population was from SEER database and the U.S. Census Bureau’s Population Estimates Program respectively, including age and calendar year at follow-up or cancer diagnosis (1990-1992, 1993-1995, 1996-1998, 1999-2001, 2002-2004, 2005-2007, 2008-2010, 2011-2015 or 2016-2020), race (white, black, or American Indian/Alaska Native, and Asian/Pacific Islander), gender (female or male), and county (counties in metropolitan areas of larger than 1 million population, counties in metropolitan areas of 250,000 to 1 million population, counties in metropolitan areas of less than 250 thousand population, nonmetropolitan counties not adjacent to a metropolitan area or nonmetropolitan counties adjacent to a metropolitan area). We classified age at follow-up, which is generally associated with risk of CVD mortality^19^, into 30-34, every 5 years, or older than 80 years, to adjust for the confounding impact of age. We also selected information on clinical Covariates from SEER for lung cancer patients, including laterality, tumor stage, histology, grade, surgery, radiotherapy, and chemotherapy. Missing data was defined as unknown. The baseline characteristics of patients and the general population were displayed in Table 1.

**Table 1.**
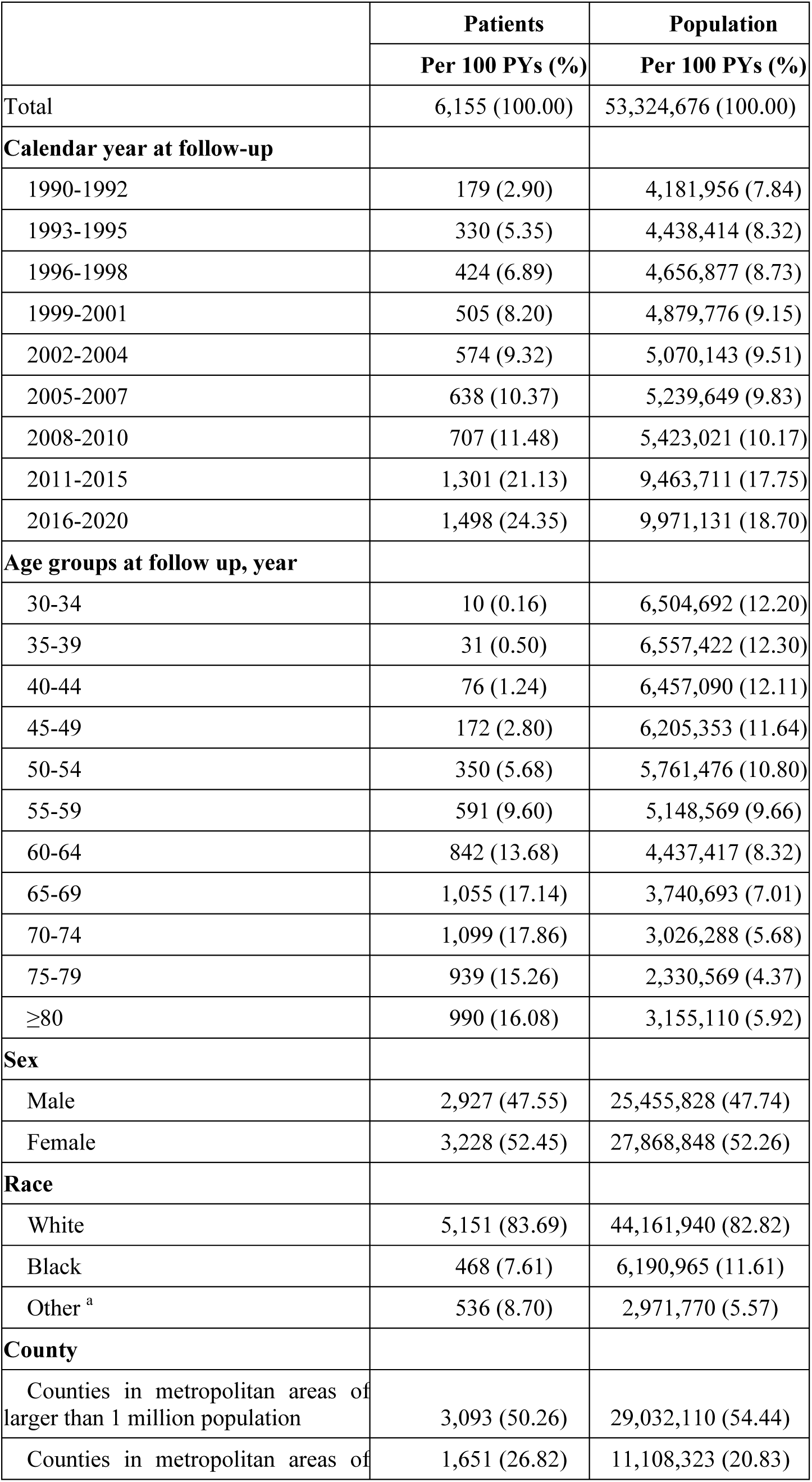
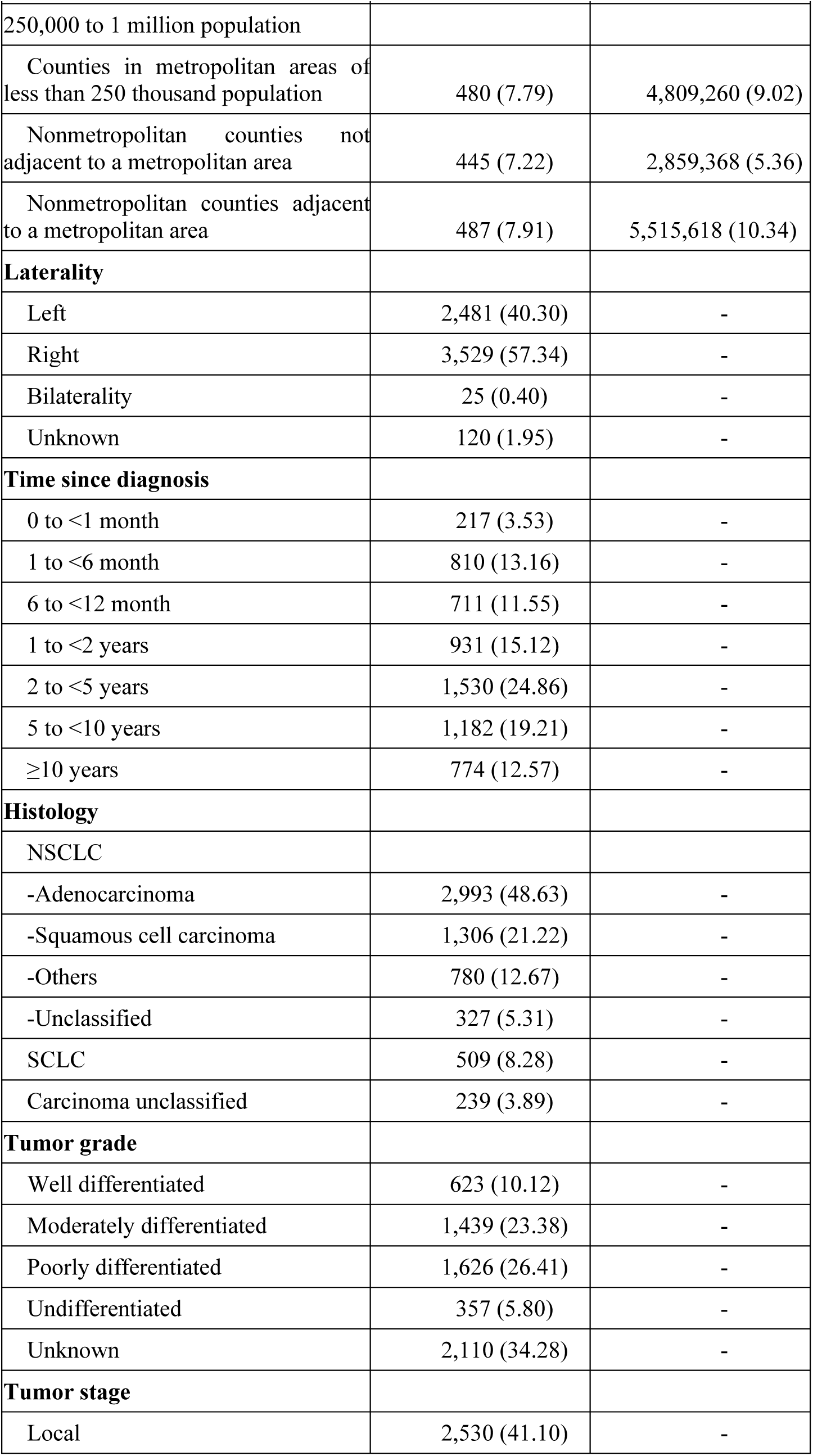
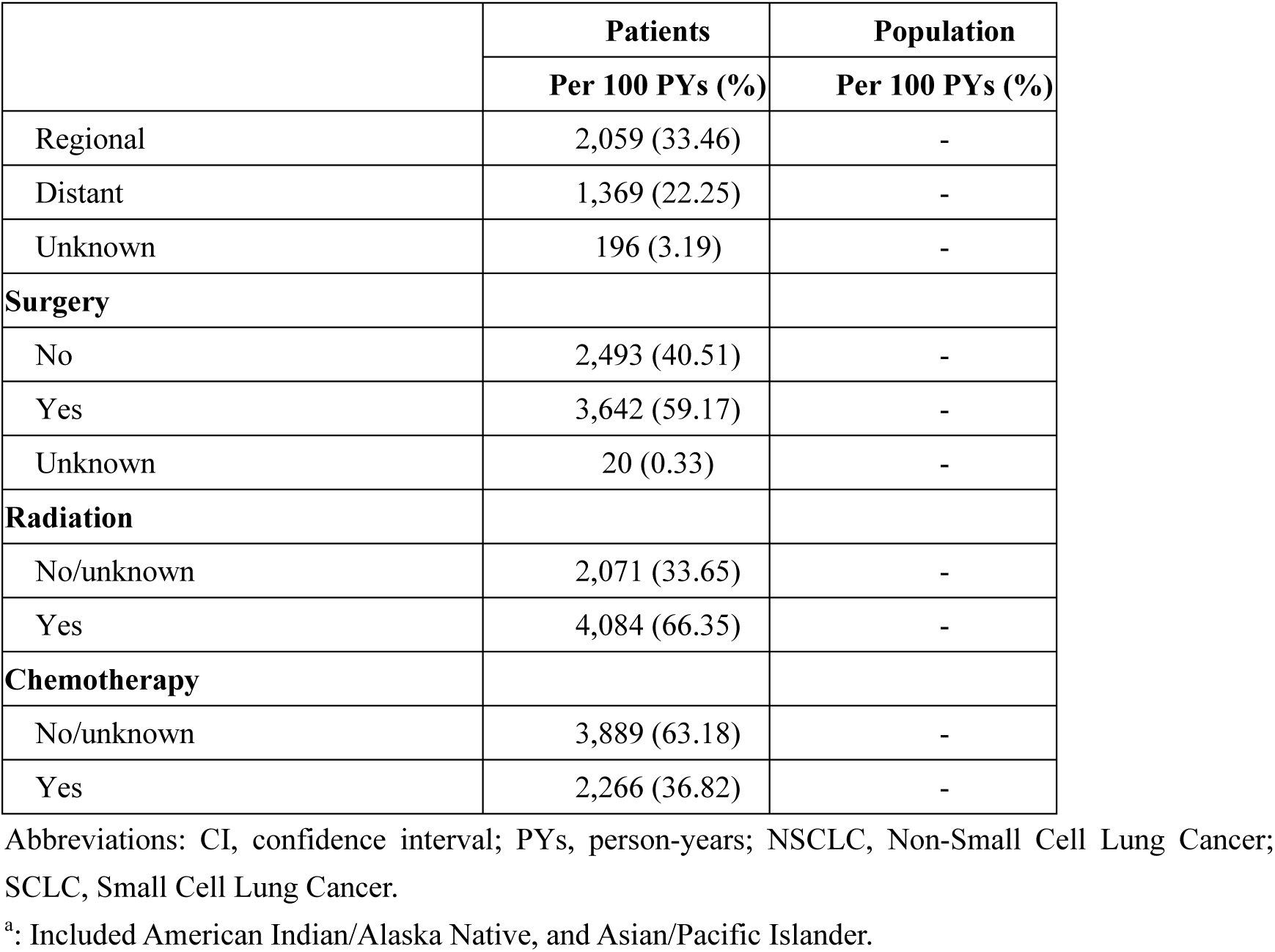
Baseline <u>characteristics</u> of lung cancer patients and the U.S. female population: a population-based cohort study in the U.S., 1990-2020.

|  | **Patients** | **Population** |
| --- | --- | --- |
|  | **Per 100 PYs (%)** | **Per 100 PYs (%)** |
| Total | 6,155 (100.00) | 53,324,676 (100.00) |
| **Calendar year at follow-up** |  |  |
| 1990-1992 | 179 (2.90) | 4,181,956 (7.84) |
| 1993-1995 | 330 (5.35) | 4,438,414 (8.32) |
| 1996-1998 | 424 (6.89) | 4,656,877 (8.73) |
| 1999-2001 | 505 (8.20) | 4,879,776 (9.15) |
| 2002-2004 | 574 (9.32) | 5,070,143 (9.51) |
| 2005-2007 | 638 (10.37) | 5,239,649 (9.83) |
| 2008-2010 | 707 (11.48) | 5,423,021 (10.17) |
| 2011-2015 | 1,301 (21.13) | 9,463,711 (17.75) |
| 2016-2020 | 1,498 (24.35) | 9,971,131 (18.70) |
| **Age groups at follow up, year** |  |  |
| 30-34 | 10 (0.16) | 6,504,692 (12.20) |
| 35-39 | 31 (0.50) | 6,557,422 (12.30) |
| 40-44 | 76 (1.24) | 6,457,090 (12.11) |
| 45-49 | 172 (2.80) | 6,205,353 (11.64) |
| 50-54 | 350 (5.68) | 5,761,476 (10.80) |
| 55-59 | 591 (9.60) | 5,148,569 (9.66) |
| 60-64 | 842 (13.68) | 4,437,417 (8.32) |
| 65-69 | 1,055 (17.14) | 3,740,693 (7.01) |
| 70-74 | 1,099 (17.86) | 3,026,288 (5.68) |
| 75-79 | 939 (15.26) | 2,330,569 (4.37) |
| ≥80 | 990 (16.08) | 3,155,110 (5.92) |
| **Sex** |  |  |
| Male | 2,927 (47.55) | 25,455,828 (47.74) |
| Female | 3,228 (52.45) | 27,868,848 (52.26) |
| **Race** |  |  |
| White | 5,151 (83.69) | 44,161,940 (82.82) |
| Black | 468 (7.61) | 6,190,965 (11.61) |
| Other <sup>a</sup> | 536 (8.70) | 2,971,770 (5.57) |
| **County** |  |  |
| Counties in metropolitan areas of larger than 1 million population | 3,093 (50.26) | 29,032,110 (54.44) |
| Counties in metropolitan areas of | 1,651 (26.82) | 11,108,323 (20.83) |

|  | Patients | Population |
| --- | --- | --- |
|  | Per 100 PYs (%) | Per 100 PYs (%) |
| 250,000 to 1 million population |  |  |
| Counties in metropolitan areas of less than 250 thousand population | 480 (7.79) | 4,809,260 (9.02) |
| Nonmetropolitan counties not adjacent to a metropolitan area | 445 (7.22) | 2,859,368 (5.36) |
| Nonmetropolitan counties adjacent to a metropolitan area | 487 (7.91) | 5,515,618 (10.34) |
| **Laterality** |  |  |
| Left | 2,481 (40.30) | - |
| Right | 3,529 (57.34) | - |
| Bilaterality | 25 (0.40) | - |
| Unknown | 120 (1.95) | - |
| **Time since diagnosis** |  |  |
| 0 to <1 month | 217 (3.53) | - |
| 1 to <6 month | 810 (13.16) | - |
| 6 to <12 month | 711 (11.55) | - |
| 1 to <2 years | 931 (15.12) | - |
| 2 to <5 years | 1,530 (24.86) | - |
| 5 to <10 years | 1,182 (19.21) | - |
| ≥10 years | 774 (12.57) | - |
| **Histology** |  |  |
| NSCLC |  |  |
| -Adenocarcinoma | 2,993 (48.63) | - |
| -Squamous cell carcinoma | 1,306 (21.22) | - |
| -Others | 780 (12.67) | - |
| -Unclassified | 327 (5.31) | - |
| SCLC | 509 (8.28) | - |
| Carcinoma unclassified | 239 (3.89) | - |
| **Tumor grade** |  |  |
| Well differentiated | 623 (10.12) | - |
| Moderately differentiated | 1,439 (23.38) | - |
| Poorly differentiated | 1,626 (26.41) | - |
| Undifferentiated | 357 (5.80) | - |
| Unknown | 2,110 (34.28) | - |
| **Tumor stage** |  |  |
| Local | 2,530 (41.10) | - |
|  | Per 100 PYs (%) | Per 100 PYs (%) |
| Regional | 2,059 (33.46) | - |
| Distant | 1,369 (22.25) | - |
| Unknown | 196 (3.19) | - |
| **Surgery** |  |  |
| No | 2,493 (40.51) | - |
| Yes | 3,642 (59.17) | - |
| Unknown | 20 (0.33) | - |
| **Radiation** |  |  |
| No/unknown | 2,071 (33.65) | - |
| Yes | 4,084 (66.35) | - |
| **Chemotherapy** |  |  |
| No/unknown | 3,889 (63.18) | - |
| Yes | 2,266 (36.82) | - |

Abbreviations: CI, confidence interval; PYs, person-years; NSCLC, Non-Small Cell Lung Cancer; SCLC, Small Cell Lung Cancer.
<sup>a</sup>: Included American Indian/Alaska Native, and Asian/Pacific Islander.

### Statistical analysis

Using Poisson regression, we calculated the incidence rate ratios (IRRs) and 95% confidence intervals (95% CIs) of CVD deaths among patients relative to the general population controlling for age at follow-up, race, gender, county, and calendar year at follow-up. We described the cumulative mortality rate for lung cancer patients by age at follow-up 80 years old. We estimated IRRs of death owe to diseases of the heart, cerebrovascular diseases, and other CVD in subgroups. We also assessed IRRs by follow-up time since cancer diagnosis. We conducted subsequently subgroup analyses in patients at age of follow-up 30-79 years old, on account of the obviously increased risk in this age group.

STATA (version 16.0; Stata Corporation) was used to calculate statistical analyses. P<0.05 shows the significant difference.

## Results

### Survival characteristics

Survival data of lung cancer patients and general population were obtained from the SEER database and U.S. Census Bureau’s Population Estimates Program between 1990 and 2020, respectively. 278,418 lung cancer patients were diagnosed during 1990-2020 with 12,584 CVD deaths (mortality rate: 2.04 per 100 person-years) and a median follow-up of 9 months (interquartile range, 3-27 months). 26,948,910 CVD deaths (mortality rate: 0.51 per 100 person-years) were identified in the general population.

### Age at follow-up

The cumulative mortality rate of CVD among lung cancer survivors at age of follow-up over 80 years was obviously higher than those 30-79 after a decade from diagnosis (6.73% vs. 3.91%; Figure 1).

**Figure 1.**
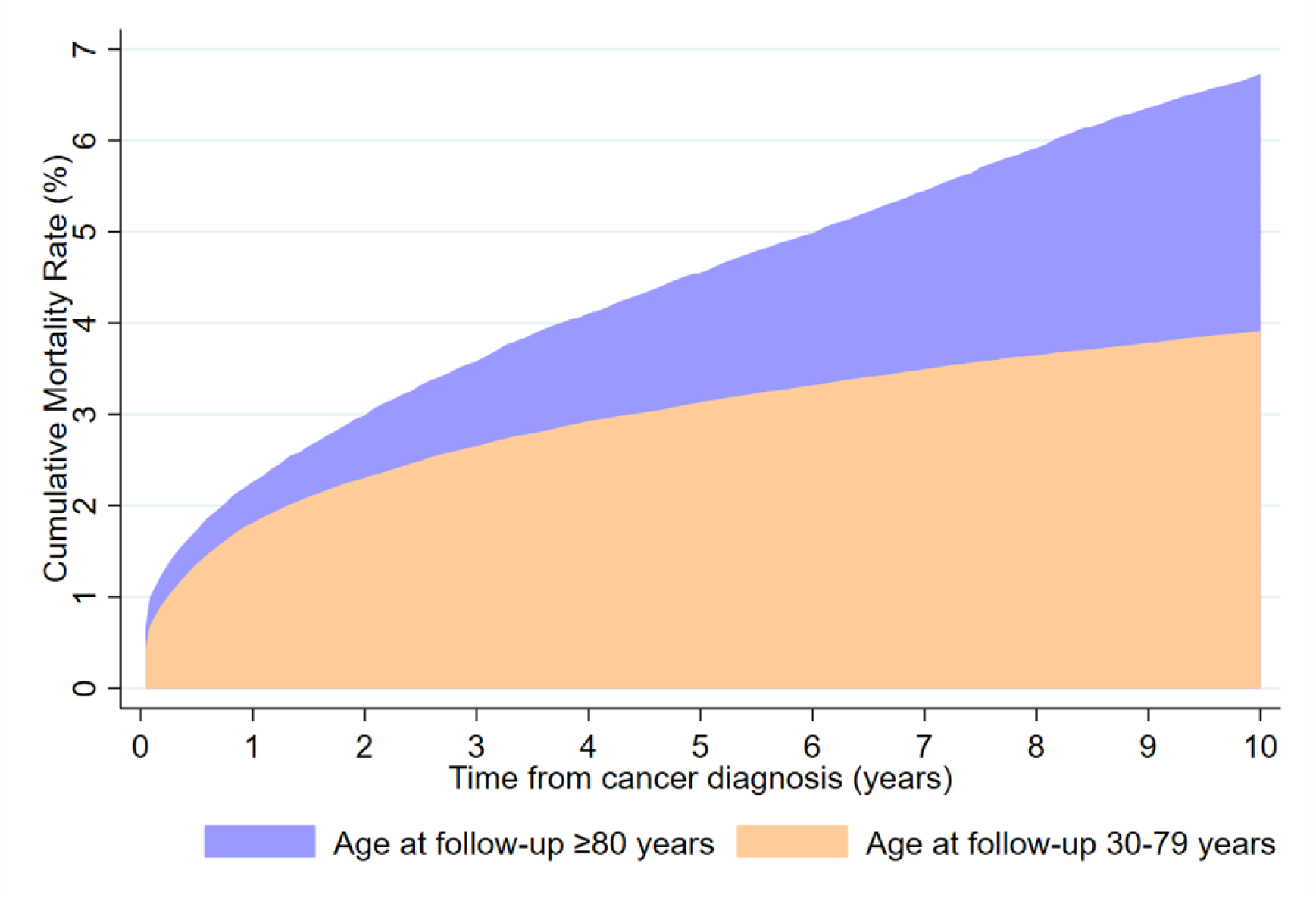
Cumulative mortality rates of cardiovascular death by age at follow-up from lung cancer diagnosis to 10 years.

Overall, lung cancer patients had a higher incidence rate ratio (IRR)of CVD mortality compared with the general population [IRR 1.74, 95% confidence interval (CI) 1.71-1.77; Table 2]. Furthermore, patients at age of follow-up 30-79 years old were correlated with a more significantly increased risk of CVD mortality (IRR 2.61, 95% CI 2.55-2.66; Table 2) and this correlation was most pronounced among those with 30-34 years old (IRR 48.93, 95% CI 21.98-108.92; Table 2).

**Table 2.**
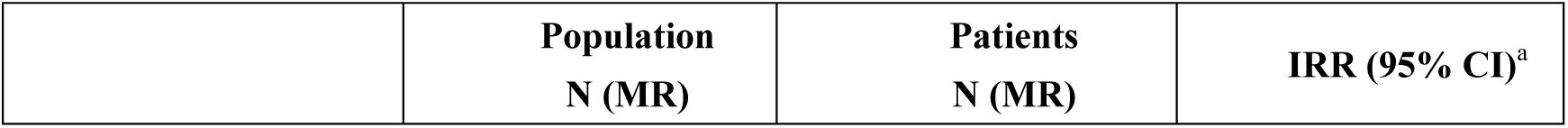
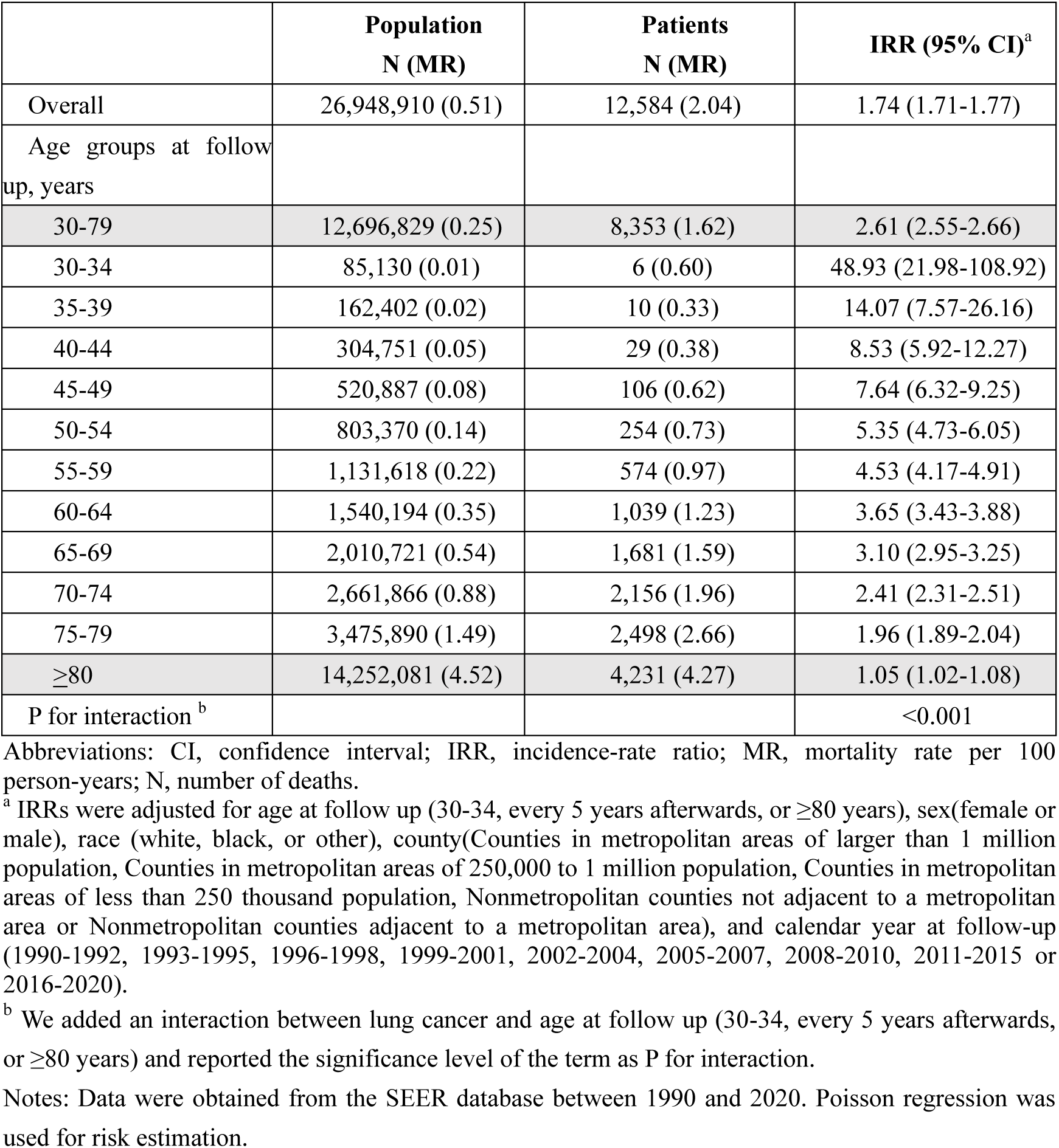
Adjusted incidence rate ratios (IRRs) for cardiovascular mortality among lung cancer patients compared to the general population, stratified by age group: a population-based cohort study in the U.S., 1990-2020.

|  | **Population**<br>**N (MR)** | **Patients**<br>**N (MR)** | **IRR (95% CI)<sup>a</sup>** |
| --- | --- | --- | --- |
| Overall | 26,948,910 (0.51) | 12,584 (2.04) | 1.74 (1.71-1.77) |
| Age groups at follow up, years |  |  |  |
| 30-79 | 12,696,829 (0.25) | 8,353 (1.62) | 2.61 (2.55-2.66) |
| 30-34 | 85,130 (0.01) | 6 (0.60) | 48.93 (21.98-108.92) |
| 35-39 | 162,402 (0.02) | 10 (0.33) | 14.07 (7.57-26.16) |
| 40-44 | 304,751 (0.05) | 29 (0.38) | 8.53 (5.92-12.27) |
| 45-49 | 520,887 (0.08) | 106 (0.62) | 7.64 (6.32-9.25) |
| 50-54 | 803,370 (0.14) | 254 (0.73) | 5.35 (4.73-6.05) |
| 55-59 | 1,131,618 (0.22) | 574 (0.97) | 4.53 (4.17-4.91) |
| 60-64 | 1,540,194 (0.35) | 1,039 (1.23) | 3.65 (3.43-3.88) |
| 65-69 | 2,010,721 (0.54) | 1,681 (1.59) | 3.10 (2.95-3.25) |
| 70-74 | 2,661,866 (0.88) | 2,156 (1.96) | 2.41 (2.31-2.51) |
| 75-79 | 3,475,890 (1.49) | 2,498 (2.66) | 1.96 (1.89-2.04) |
| ≥80 | 14,252,081 (4.52) | 4,231 (4.27) | 1.05 (1.02-1.08) |
| P for interaction<sup>b</sup> |  |  | <0.001 |

Abbreviations: CI, confidence interval; IRR, incidence-rate ratio; MR, mortality rate per 100 person-years; N, number of deaths.
<sup>a</sup> IRRs were adjusted for age at follow up (30-34, every 5 years afterwards, or ≥80 years), sex(female or male), race (white, black, or other), county(Counties in metropolitan areas of larger than 1 million population, Counties in metropolitan areas of 250,000 to 1 million population, Counties in metropolitan areas of less than 250 thousand population, Nonmetropolitan counties not adjacent to a metropolitan area or Nonmetropolitan counties adjacent to a metropolitan area), and calendar year at follow-up (1990-1992, 1993-1995, 1996-1998, 1999-2001, 2002-2004, 2005-2007, 2008-2010, 2011-2015 or 2016-2020).
<sup>b</sup> We added an interaction between lung cancer and age at follow up (30-34, every 5 years afterwards, or ≥80 years) and reported the significance level of the term as P for interaction.
Notes: Data were obtained from the SEER database between 1990 and 2020. Poisson regression was used for risk estimation.

**Table 3.** Incidence rate ratios (IRRs) of cardiovascular mortality in lung cancer patients compared to the general population by subgroup: a population-based study in the U.S., 1990-2020.

|  | Population<br>N (MR) | Patients<br>N (MR) | IRR (95% CI)<sup>a</sup> |
| --- | --- | --- | --- |
| **Diseases of heart** |  |  |  |
| Overall | 20,669,892 (0.39) | 9,834 (1.60) | 1.79 (1.75-1.82) |
| Age at follow-up |  |  |  |
| 30-79 years | 10,027,554 (0.20) | 6,555 (1.27) | 2.62 (2.56-2.69) |
| ≥80 years | 10,642,337 (3.37) | 3,279 (3.31) | 1.09 (1.05-1.13) |
| **Cerebrovascular diseases** |  |  |  |
| Overall | 4,523,101 (0.09) | 1,882 (0.31) | 1.52 (1.45-1.59) |
| Age at follow-up |  |  |  |
| 30-79 years | 1,869,634 (0.04) | 1,207 (0.23) | 2.44 (2.31-2.58) |
| ≥80 years | 2,653,467 (0.84) | 675 (0.68) | 0.91 (0.84-0.98) |
| **Other cardiovascular diseases** |  |  |  |
| Overall | 1,743,190 (0.03) | 868 (0.14) | 1.78 (1.67-1.90) |
| Age at follow-up |  |  |  |
| 30-79 years | 788,329 (0.02) | 591 (0.11) | 2.80 (2.59-3.04) |
| ≥80 years | 954,861 (0.30) | 277 (0.28) | 1.00 (0.89-1.12) |

Abbreviations: CI, confidence interval; IRR, incidence-rate ratio; MR, mortality rate per 100 person-years; N, number of deaths.
<sup>a</sup> IRRs were adjusted for age at follow up (30-34, every 5 years afterwards, or ≥80 years), sex(female or male), race (white, black, or other), county(Counties in metropolitan areas of larger than 1 million population, Counties in metropolitan areas of 250,000 to 1 million population, Counties in metropolitan areas of less than 250 thousand population, Nonmetropolitan counties not adjacent to a metropolitan area or Nonmetropolitan counties adjacent to a metropolitan area), and calendar year at follow-up (1990-1992, 1993-1995, 1996-1998, 1999-2001, 2002-2004, 2005-2007, 2008-2010, 2011-2015 or 2016-2020).
Notes: Data were obtained from the SEER database between 1990 and 2020. Poisson regression was used for risk estimation.

In the subgroup analysis, a greater risk association in lung cancer patients at age of follow-up 30-79 years was found for deaths from heart of diseases, cerebrovascular, as well as other CVD relative to general

### Demographic and clinical characteristics

We performed subsequent analyses by restricting patients in 30-79 years old, Since the increased risk was profound at this age stage. A higher magnitude associations was found among lung cancer patients with female gender (IRR 2.82, 95% CI 2.72-2.92), who lived counties in metropolitan areas of 250,000 to 1 million population (IRR 2.94, 95% CI 2.83-3.06), early calendar year at follow up(1990-1992, IRR 3.75, 95% CI 3.46-4.07), Small Cell Lung Cancer (SCLC) (IRR 3.53, 95% CI 3.32-3.77), bilaterality side carcinoma (IRR 3.82, 95% CI 2.91-5.02), distant stage (IRR 4.03, 95% CI 3.88-4.19), undifferentiated tumor grade (IRRs 3.12, 95% CI 2.89-3.37), or who didn’t receive chemotherapy, radiation or surgery (IRR 2.66-3.90) (Table 4; Table 5). Besides, our findings also found that lung cancer patients who received chemotherapy (IRR 2.50, 95% CI 2.41-2.60) or radiotherapy (IRR 2.44, 95% CI 2.37-2.51) exhibited significantly higher risks of cardiovascular mortality compared to general population (Table 5).

**Table 4.**
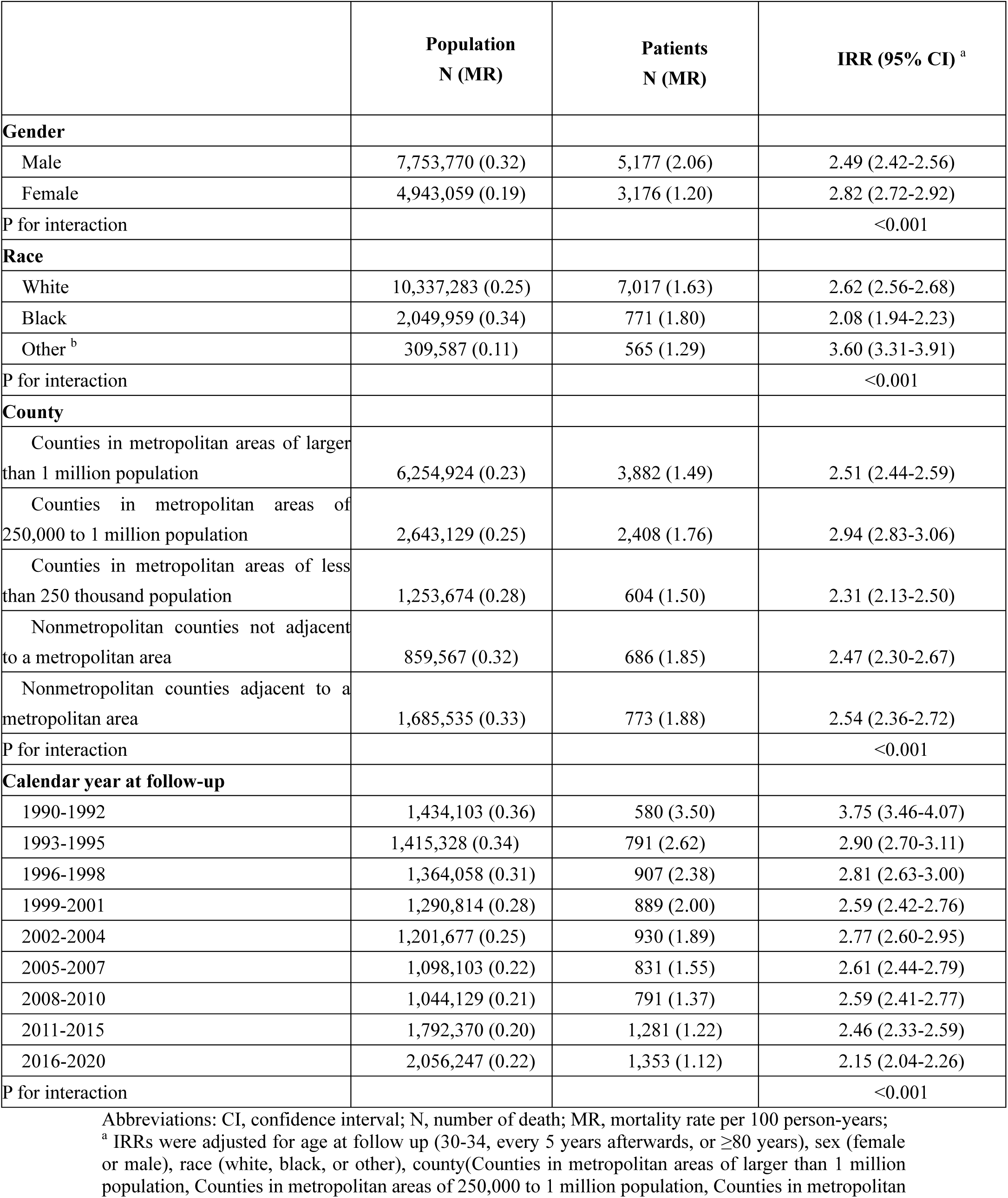
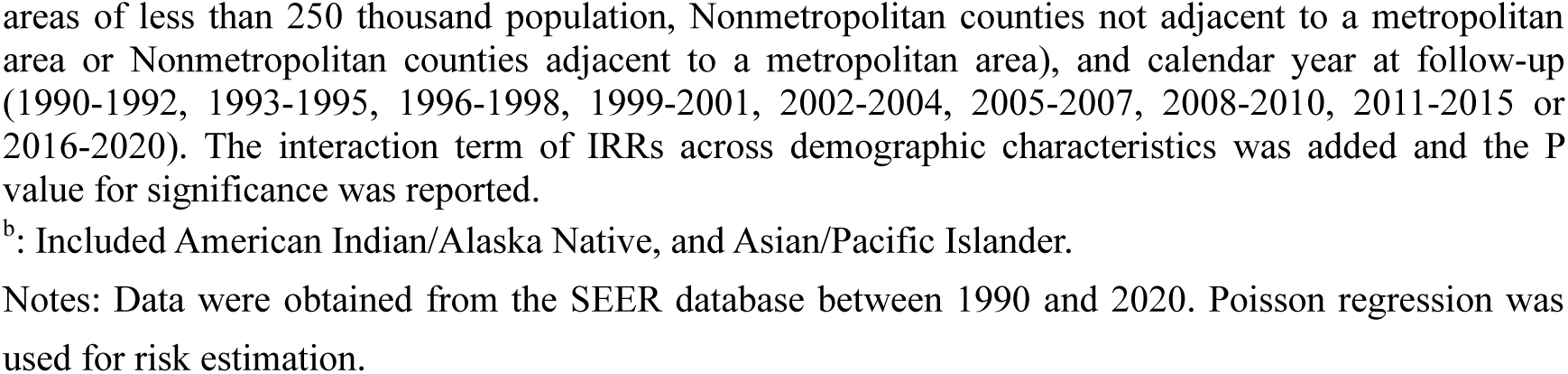
Incidence-rate ratios (IRRs) of cardiovascular mortality at age of follow up 30-79 years in lung cancer patients compared to the general population, stratified by demographic characteristics: a population-based study in the U.S., 1990-2020.

**Table 5.** Incidence-rate ratios (IRRs) of cardiovascular mortality at age of follow up 30-79 years in lung cancer patients compared to the general population, by clinical characteristics: a population-based study in the U.S., 1990-2020.

|  | **Patients**<br>**N (MR)** | **IRR (95% CI) <sup>a</sup>** |
| --- | --- | --- |
| **Laterality** |  |  |
| Left | 5315 (2.14) | 2.63 (2.55-2.72) |
| Right | 6826 (1.93) | 2.51 (2.44-2.59) |
| Bilaterality | 71 (2.88) | 3.82 (2.91-5.02) |
| P for difference |  | <0.001 |
| **Histology** |  |  |
| NSCLC | 10577 (1.96) | 2.45 (2.39-2.51) |
| -Adenocarcinoma (NSCLC) | 5123 (1.71) | 2.23 (2.16-2.31) |
| -Squamous cell carcinoma (NSCLC) | 3506 (2.68) | 2.73 (2.63-2.85) |
| -Others (NSCLC) | 1117 (1.43) | 2.00 (1.86-2.16) |
| -Unclassified (NSCLC) | 831 (2.54) | 3.89 (3.59-4.22) |
| SCLC | 1163 (2.28) | 3.53 (3.32-3.77) |
| Carcinoma unclassified | 844 (3.52) | 3.84 (3.54-4.17) |
| P for difference |  | <0.001 |
| **Tumor stage** |  |  |
| Local | 4504 (1.78) | 1.92 (1.85-2.00) |
| Regional | 3938 (1.91) | 2.48 (2.39-2.57) |
| Distant | 3469 (2.53) | 4.03 (3.88-4.19) |
| Unstaged | 673 (3.43) | 3.42 (3.11-3.77) |
| P for difference |  | <0.001 |
| **Tumor grade** |  |  |
| Well differentiated | 866 (1.39) | 1.57 (1.43-1.73) |
| Moderately differentiated | 2457 (1.71) | 2.01 (1.91-2.11) |
| Poorly differentiated | 3451 (2.12) | 2.57 (2.47-2.68) |
| Undifferentiated | 849 (2.38) | 3.12 (2.89-3.37) |
| P for difference |  | <0.001 |
| **Chemotherapy** |  |  |
| No/unknown | 9281 (2.39) | 2.66 (2.59-2.73) |
| Yes | 3303 (1.46) | 2.50 (2.41-2.60) |
| P for difference |  | <0.001 |
| **Radiotherapy** |  |  |
| No/unknown | 4357 (2.10) | 2.93 (2.83-3.03) |
| Yes | 8227 (2.01) | 2.44 (2.37-2.51) |
| P for difference |  | <0.001 |
| **Surgery** |  |  |
| No/unknown | 7274 (2.89) | 3.90 (3.80-4.01) |
| Yes | 5310 (1.46) | 1.71 (1.66-1.77) |
| P for difference |  | <0.001 |

Abbreviations: CI, confidence interval; PYs, person-years; N, number of death; MR, mortality rate per 100 person-years; NSCLC, Non-Small Cell Lung Cancer; SCLC, Small Cell Lung Cancer.
<sup>a</sup> IRRs were adjusted for age at follow up (30-34, every 5 years afterwards, or ≥80 years), sex (female or male), race (white, black, or other), county (Counties in metropolitan areas of larger than 1 million population, Counties in metropolitan areas of 250,000 to 1 million population, Counties in metropolitan areas of less than 250 thousand population, Nonmetropolitan counties not adjacent to a metropolitan area or Nonmetropolitan counties adjacent to a metropolitan area), and calendar year at follow-up (1990-1992, 1993-1995, 1996-1998, 1999-2001, 2002-2004, 2005-2007, 2008-2010, 2011-2015 or 2016-2020). These IRRs were estimated by comparing clinical characteristics among lung cancer patients with the general population. The difference of IRRs across characteristics was performed using Wald test and the P value for significance was reported.
Note: Population was regarded as reference [ N (MR):26,948,910 (0.51) ] in every estimate. Data were obtained from the SEER database between 1990 and 2020. Poisson regression was used for risk estimation.

### Follow-up time since lung cancer diagnosis

The risks of CVD mortality were greatest among lung cancer patients at the age of follow up 30-79 and ≥80 years, respectively, during first month after lung cancer diagnosis (IRR 12.08, 95% CI 11.49-12.70 and IRR 4.03, 95% CI 3.7-4.39; Figure 2 and statistical values in Table S2), compared with the population.

**Figure 2.**
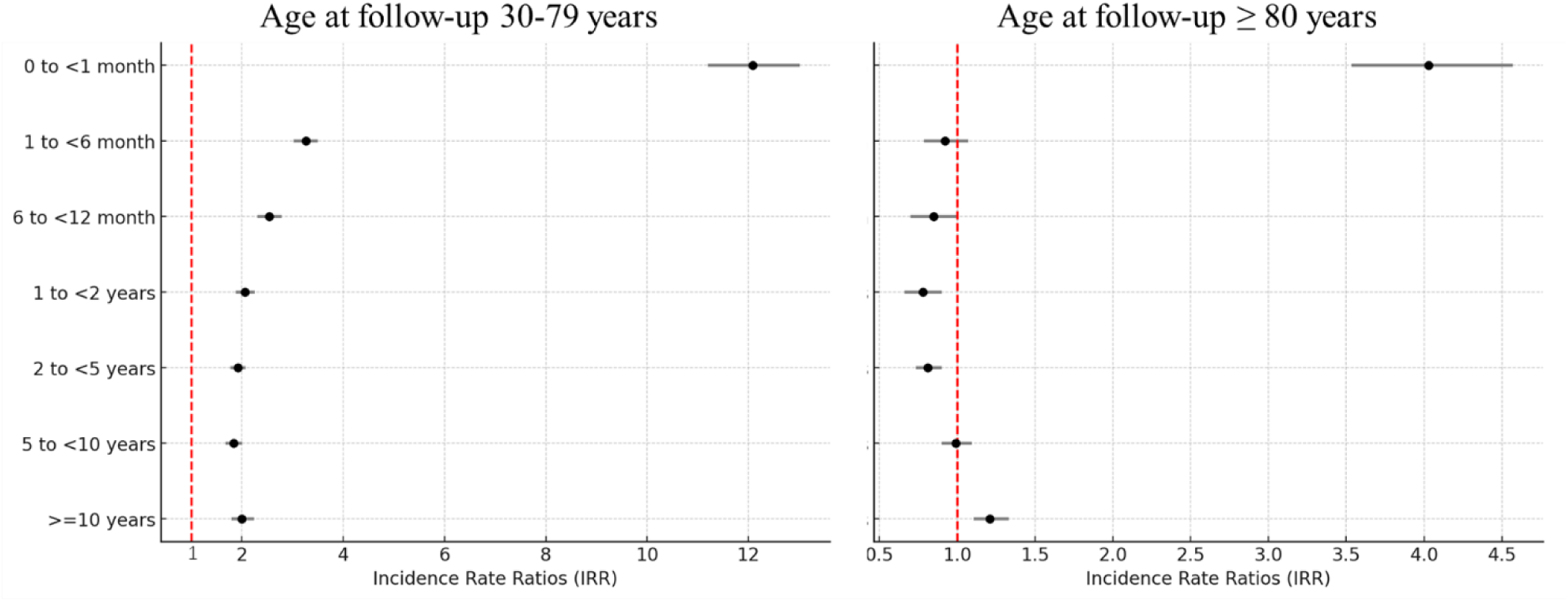
Incidence rate ratios (IRRs) of cardiovascular mortality by time since cancer diagnosis, comparing lung cancer patients with the general population: a population-based study in the U.S., 1990-2020.

## Discussion

The population-based cohort study demonstrates that lung cancer patients face a significantly higher risk of cardiovascular mortality compared to the general population. The risk peaks during the first month after lung cancer diagnosis and is particularly pronounced in younger patients.A study reported that patients with lung cancer had standardized mortality ratio (SMRs) of 7–14 fatal heart disease in the first year after diagnosis and this trend remained elevated throughout all follow-up times^20^. Our findings extensively indicate that early diagnosis of lung cancer is associated with a significantly increased risk of CVD mortality, particularly within the first month post-diagnosis. During this critical period, patients exhibit greatest increased risk of CVD mortality in younger (ages 30–79; IRR 12.08, 95% CI 11.49-12.70) and older (ages ≥ 80; IRR 4.03, 95% CI 3.70-4.39) groups, respectively, compared to the general population. Several factors likely contribute to this early and heightened cardiovascular risk. Firstly, being diagnosed with lung cancer, a life-threatening illness, can induce significant psychological stress, which is linked to adverse cardiovascular outcomes^17^. Stress-related mechanisms include increased sympathetic nervous system activity and higher levels of circulating catecholamines, which can precipitate cardiovascular events such as myocardial infarction and stroke^21,22^.

Secondly, the initiation of cancer treatments may exacerbate pre-existing cardiovascular conditions. Surgery induces significant perioperative stress, increasing the risk of thromboembolism and arrhythmias^23^. Similarly, chemotherapy (tyrosine kinase inhibitors) and immunotherapy are associated with acute myocardial injury, while radiation therapy can induce vascular inflammation and fibrosis^12,24–26^. Thirdly, systemic inflammation induced by both cancer progression and its treatment play a critical role. Pro-inflammatory cytokines, such as IL-6 and TNF-α, are significantly elevated in lung cancer patients and contribute to endothelial dysfunction, plaque instability, and increased coagulation, all of which heighten cardiovascular risk^27,28^.These findings underscore the importance of close cardiovascular monitoring after lung cancer diagnosis, particularly for younger patients. Tailored cardio-oncology care strategies, including baseline cardiovascular risk assessment and early intervention, could significantly mitigate this heightened risk and improve patient outcomes. In addition, temporal analysis reveals a decline in cardiovascular mortality risk over the study period (1990-2020), particularly from 2010 onward, coinciding with the introduction of immunotherapy and advanced chemotherapy protocols. These findings highlight the impact of evolving treatment strategies on cardiovascular outcomes^29^.

Radiation therapy may elevate the risk of CVD mortality by triggering acute inflammatory cascades, which can lead to myocardial fibrosis and subsequent damage to cardiac muscle and surrounding vasculature^25^. Chemotherapy, particularly tyrosine kinase inhibitors, also leads to a higher risk of CVD mortality due to injury of the circulation system^11^. Our findings of an increased risk of CVD mortality in lung cancer patients who received radiation/chemotherapy validate this association. Although female patients with lung cancer have a lower absolute risk of CVD-specific death relative to male ones^3^, they exhibit a relatively higher risk when compared to the general female population. This may be due to a higher frequency of cardiotoxicity observed in women undergoing cancer treatment^25,30^.Cardiotoxicity from lung cancer treatment extends beyond left ventricular dysfunction, which is often a late-stage manifestation of cardiac damage. Emerging evidence highlights the utility of more sensitive metrics for early detection. Techniques such as myocardial strain analysis via echocardiography and biomarkers like cardiac troponins and BNP offer valuable insights into subclinical cardiac injury^31^. These measures can detect myocardial damage long before symptoms appear or left ventricular ejection fraction declines, thus allowing for timely alleviating the cumulative cardiovascular burden of lung cancer treatment and reducing long-term complications. In addition to treatment-related cardiotoxicity, gender differences in CVD mortality among lung cancer patients are also influenced by biological and sociobehavioral factors. Biologically, hormonal factors such as the cardioprotective effects of estrogen might partially explain the lower absolute CVD mortality risk in women compared to men^32^. In male patients, coronary artery disease primarily affects the epicardial coronary arteries, whereas in female patients, the microvascular circulation is most significantly impacted, which may increase women’s susceptibility to cardiotoxic effects^33^. Sociobehavioral factors also play a role. In female patients, tobacco use, obesity, type 2 diabetes mellitus, depression, and psychosocial stress have a more powerful impact on CVD compared to male patients^33^. Addressing these disparities requires a gender-sensitive approach to cardio-oncology care, including proactive cardiovascular monitoring and tailored interventions for women undergoing cancer treatment. Patients with poorly differentiated tumors or bilateral lung cancer face a higher risk of CVD mortality due to complex treatment methods^24^, heavy tumor burden, systemic inflammation and metabolic disorders^34^. Our analysis suggests advanced-stage lung cancer correlates with higher CVD mortality, highlighting the importance of cardiovascular management, particularly for younger patients and those who have not undergone treatment. Moreover, our findings demonstrate SCLC patients exhibit a significantly higher cardiovascular mortality risk (IRR 3.53, 95% CI: 3.32–3.77) than both the general population and NSCLC patients. The aggressive nature of SCLC leads to intense treatment regimens, including high-dose chemotherapy and thoracic radiotherapy, both of which are known to have cardiotoxic effects^35^. Rapid tumor progression and higher baseline systemic inflammation in SCLC patients may further exacerbate cardiovascular mortality risks^36^.

Aging is a significant factor in both CVD and lung cancer risk^5,9^. Our findings indicate that CVD mortality rates increase with age in both lung cancer patients and the general population. Our findings indicate that absolute CVD mortality rates increase with age in both lung cancer patients and the general population. Interestingly, in comparison with general population, our study shows a dramatic increase in relative CVD mortality risk among patients aged 30–34, while those aged 80 and older experience a milder increase. This aligns with recent findings suggesting that younger cancer survivors face a disproportionately higher risk of heart disease-related death compared to their age-matched counterparts in the general population^5^.

Clinically, this study provides actionable insights for improving patient care. It underscores the necessity of integrating cardiovascular risk assessment and management into lung cancer treatment protocols, particularly during the first month post-diagnosis and for younger or high-risk subgroups. By identifying specific demographic and clinical factors associated with increased CVD mortality, this research offers a roadmap for tailoring cardio-oncology interventions to enhance patient outcomes. In addition, this study serves as a call to action for future research. It highlights the need to explore the mechanisms underlying age- and temporal-specific differences in cardiotoxicity, investigate the long-term impact of cancer therapies on cardiovascular health, and develop preventive strategies that balance cancer treatment efficacy with cardiovascular safety.

## Limitations

Some limitations should be noted in this study. First, available information on general population is none cancer-free, contributing to underrate the real risk of CVD mortality. Second, the lack of data on comorbid conditions such as hypertension and diabetes is a limitation of this study. These conditions are known to exacerbate cardiovascular risks, particularly in lung cancer patients undergoing aggressive treatment. Still and all, the significantly elevated risk of CVD mortality within one month after cancer diagnosis alleviates this concern. Last, though the distribution characteristics of lung cancer patients and population in age at follow-up is unbalanced, we still investigated the analyzable association by age between the two groups.

## Conclusions

Our results show lung cancer patients are at increased risk of CVD mortality, compared with the general population. Our findings underscore the potential care for monitoring CVD among lung cancer patients especially who were young or with advanced stage. The greatest risk of CVD mortality during the first month post lung cancer diagnosis may need the integration of tailored CVD care strategies.

## Data availability statement

Publicly available datasets could be accessed from Surveillance, Epidemiology, and End Results Program (SEER) database.

## Author contributions

Conception and design: Chengshi Wang, Zhu Wang, and Ye Yang. Collection and assembly of data: Jing Yang and Songbo Zhang. Data analysis and interpretation: Purong Zhang, Chengshi Wang, and Jing Yang. Manuscript writing: All authors. All authors contributed to the article and approved the submitted version.

## Publisher’s note

All claims expressed in this article are solely those of the authors and do not necessarily represent those of their affiliated organizations, or those of the publisher, the editors and the reviewers. Any product that may be evaluated in this article, or claim that may be made by its manufacturer, is not guaranteed or endorsed by the publisher.

